# Comparative Effectiveness of Recommended and Delayed Dosing Schedules for Rotavirus Vaccine: Target Trial Emulation

**DOI:** 10.64898/2026.07.13.26357904

**Authors:** Shae Gantt, Toshiaki Komura, Elizabeth T Rogawski McQuade, Kayoko Shioda

**Affiliations:** Department of Biostatistics, School of Public Health, Boston University, Boston, MA; Department of Social and Behavioral Sciences, Harvard T. H. Chan School of Public Health, Boston, MA; Department of Epidemiology, Rollins School of Public Health, Emory University, Atlanta, GA; Department of Global Health, School of Public Health, Boston University, Boston, MA; Center on Emerging Infectious Diseases, Boston University, Boston, MA

**Keywords:** causal inference, clone-censor-weight analysis, rotavirus vaccine, target trial emulation, vaccine dosing schedules, vaccine evaluation

## Abstract

**Background:** Live oral rotavirus vaccines were found to be less effective in low-income countries compared to high-income countries when using the same product and dosing schedule. We investigated whether altering dose timing may improve immune protection using a target trial emulation approach.

**Methods:** We emulated a target trial with clone-censor weighting to compare the effectiveness of the recommended 2-dose rotavirus vaccine schedule with a delayed schedule among children under two years of age in Peru and Brazil. Secondary data from the Malnutrition and Enteric Disease Study (MAL-ED) birth cohort (2009-2014) were analyzed. Children were followed from the date of birth until the earliest occurrence of a rotavirus outcome (infection confirmed by PCR or enzyme immunoassay (EIA) or diarrhea confirmed by EIA), protocol nonadherence, loss to follow-up, or their second birthday.

**Results:** We included 154 children in Brazil and 192 in Peru. At two years of follow-up, the risk ratio (RR) for PCR-confirmed infection, using the recommended schedule as the reference, was 1.00 (95% confidence interval [CI]: 0.73-1.37) in Peru and 0.85 (95% CI: 0.31-1.66) in Brazil. In Peru, the delayed schedule was associated with a higher cumulative risk of EIA-confirmed rotavirus diarrhea (RR at two years: 1.73; 95% CI: 1.02-2.77).

**Conclusions:** Delaying the two-dose rotavirus vaccine schedule did not change the cumulative risk of rotavirus infection, but the delayed schedule was associated with a higher risk of rotavirus diarrhea in Peru, where rotavirus incidence was higher.

## Introduction

Live oral rotavirus vaccines (Rotarix [RV1]; RotaTeq [RV5]; ROTAVAC; ROTASIIL) have been introduced in 136 countries as of 2025.^1^ While these vaccines have contributed to reducing the burden of rotavirus infections and diseases across the world, their effectiveness was found to be lower in low- and -middle-income countries (LMICs) compared to high-income countries when using the same vaccine product and standard dosing schedule.^2–7^ The vaccine efficacy against severe rotavirus diarrhea from Phase III trials has consistently been higher in high-income countries compared to LMICs, such as the 98% efficacy in Finland and the United States, but 49.4% in Malawi and 76.9% in South Africa, and 55.1% in India.^4,8,9^ A meta-analysis found substantially lower and more rapidly waning efficacy in high-mortality settings compared with high-income, low-mortality settings.^5^ Hypotheses of the lower efficacy of rotavirus vaccines in LMICs may include high rates of malnutrition, impaired immune response due to malnutrition and chronic inflammatory intestinal disease (environmental enteric dysfunction), and high frequency of repeated exposures due to environmental factors, such as inadequate clean water, sanitation, and hygiene.^7,10^ This reduced efficacy poses a threat to children’s health in LMICs, as diarrhoeal disease remains a leading cause of childhood malnutrition and death worldwide, with rotavirus identified as one of the leading pathogens.^11^ Additionally, the overwhelming majority of childhood mortality from diarrhoeal diseases occurred in LMICs in 2021.^12^

Vaccine dosing schedules often affect both the effectiveness and safety of various vaccines.^13–21^ For the two-dose rotavirus vaccine RV1, the recommended timing begins at 6 weeks for the first dose and 10 weeks for the second dose.^22,23^ To compare different timings of doses, a randomized controlled trial (RCT) in Dhaka, Bangladesh, compared children who received RV1 under the delayed schedule (10 weeks for the first dose and 17 weeks for the second dose) to those who did not receive RV1.^19^ The delayed schedule resulted in a vaccine efficacy of 73.5%.^19^ This was higher than the previously reported 48.3% vaccine efficacy for severe rotavirus infection under the standard schedule for RV5 in Bangladesh and Vietnam.^23^ These findings highlighted the need for real-world data to evaluate the alternative dose timings of rotavirus vaccines for diverse settings.

Therefore, we conducted a target trial emulation to compare the direct effect of the recommended and delayed dosing schedules of the 2-dose RV1 in preventing rotavirus infection and diarrhea for children under two years old in Peru and Brazil. The target trial emulation approach leverages observed variations of vaccine dose timings while emulating the key elements of a randomized controlled trial to limit biases that are common to observational studies.^24^ Through inferring causal contrasts using real-world data, we investigated whether the risk of rotavirus outcomes differed between the recommended and delayed schedules in Peru and Brazil.

## Methods

### Data set and study population

We used secondary data from the Malnutrition and Enteric Disease Study (MAL-ED), a longitudinal birth cohort that followed up children from birth to their second birthday across eight LMICs (Bangladesh, Brazil, India, Nepal, Peru, Pakistan, South Africa, and Tanzania) from 2009 to 2014.^25^ This analysis included data from Brazil and Peru as the other countries had not yet introduced rotavirus vaccines by the time of the MAL-ED study. Although South Africa had introduced the rotavirus vaccine, it was not included in this analysis as its recommended dosing schedule differed (the second dose is administered at 14 weeks rather than the 10 weeks used in other countries).^26^

In the MAL-ED study, each pair of mother and child was visited twice weekly for biospecimen collection, health assessments, and survey data. Non-diarrheal surveillance stool samples were collected monthly throughout follow-up to assess asymptomatic infection of enteric pathogens. Stool samples were also collected during each diarrheal episode, defined as three or more loose stools within 24 hours or the presence of blood in the stool. Stool samples were tested for rotavirus by RT-PCR and enzyme immunoassay (EIA). EIA was conducted for all children, while RT-PCR was conducted for only those who were followed for two years.

We extracted data on infant sex, weight, height, and the WAMI index at birth. The WAMI index is a socioeconomic status (SES) index, composed of access to improved water and sanitation, asset ownership, maternal education, and household income, ranging from 0 to 1 with lower scores indicative of lower SES. We also extracted data on rotavirus infection, diarrheal symptoms, and rotavirus vaccination dates among infants during their follow-up period. The MAL-ED study followed 208 children in Peru and 169 in Brazil.^27^ Due to missing data, our analysis included 192 children in Peru and 154 in Brazil in the target trial emulation (**Supplementary Methods**).

### Exposures and outcomes

We compared the effectiveness of two rotavirus vaccine dosing schedules, recommended and delayed, against rotavirus infection and diarrhea. The recommended schedule followed the standard 2-dose schedule, administering the first dose between 38 and 65 days after birth and the second dose between 66 and 168 days after birth, with at least 28 days between the doses.^22^ Based on observed variation in the timing of rotavirus vaccine administration in the MAL-ED data, we defined a delayed dosing schedule as the first dose administered between 66 and 104 days after birth and the second dose administered between 105 and 240 days after birth, with at least 28 days between the doses. We defined the following three rotavirus outcomes: 1) rotavirus infection as having a cycle threshold value less than 35 by RT-PCR, 2) rotavirus infection confirmed by EIA, and 3) symptomatic rotavirus diarrhea confirmed by EIA.

### Target trial emulation: clone-censor weight analysis

Using clone-censor-weight analysis, we emulated a hypothetical target trial to compare the direct effect (i.e., the effect of an individual’s assigned dosing schedule on their own risk of rotavirus outcomes, independent of population-level transmission effects) of the recommended and delayed RV1 dosing schedules (**Supplementary Table 1**). We estimated the risk of rotavirus infection and diarrhea among children under two years of age under the two schedules in Peru and Brazil. This analysis allowed us to estimate the per-protocol effect of these dosing schedules, using observational data.^24,28,29^

From the study population in each country, we created two clones of each individual, assigning one to the recommended schedule and the other to the delayed schedule (**Figure 1**). Within the target trial emulation framework, this approach aligns time zero (i.e., birth) across dosing strategies and allows individuals whose observed vaccination timing is initially consistent with multiple schedules to contribute to each relevant schedule, thereby preventing immortal time bias in per-protocol comparisons.^24,28,29^

**Figure 1:**
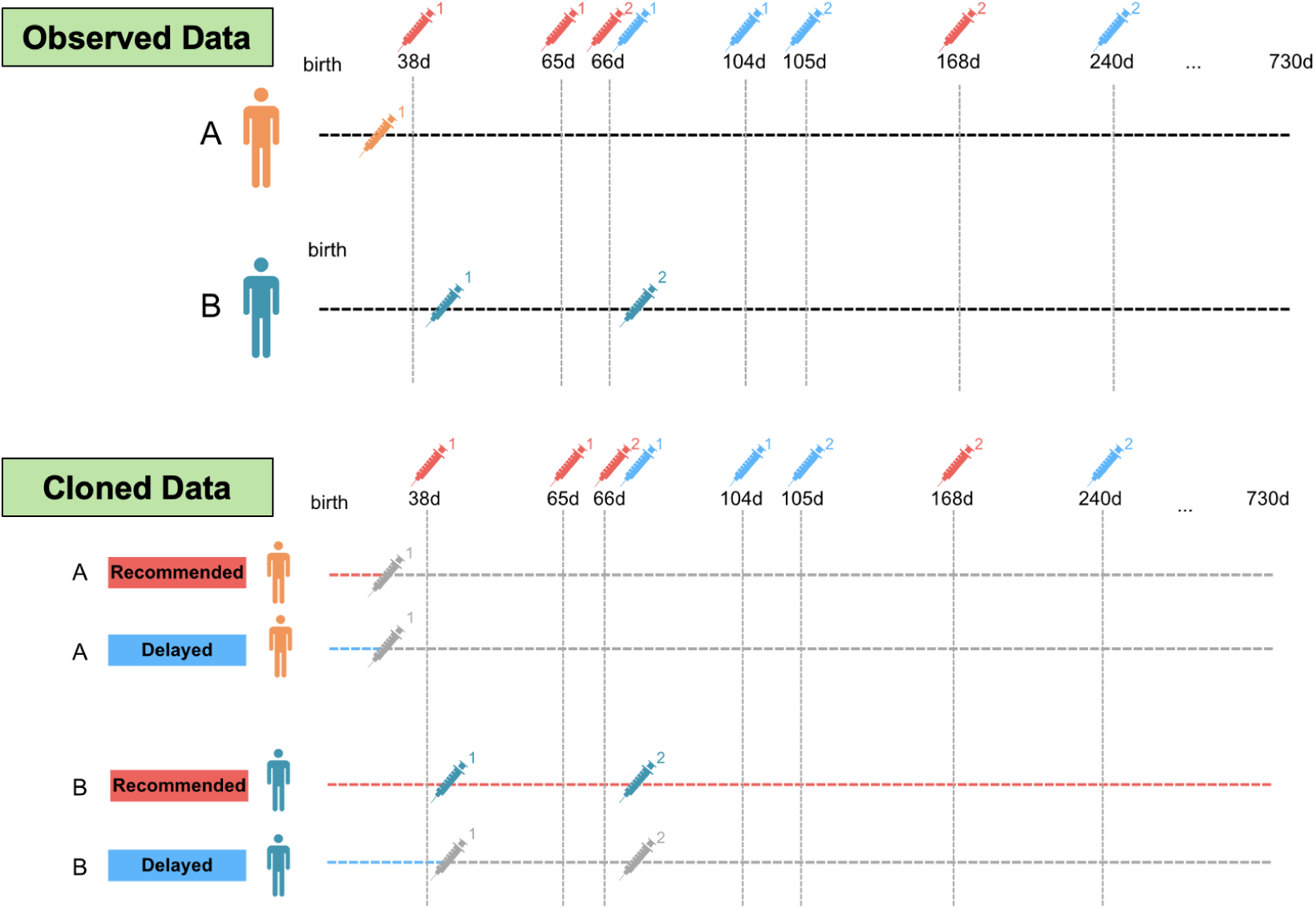
Example of follow-up in cloned dataset for two individuals over 2 years (730 days) of follow-up corresponding to the recommended and delayed dosing schedules.

Follow-up began on the child’s date of birth (index date) and continued until the first occurrence of a rotavirus outcome, protocol non-adherence, loss to follow-up, or the end of the two-year follow-up period (**Figure 1** and **Supplementary Table 2**). Non-adherence was defined as not following the assigned dosing schedule, including receipt of a dose outside the defined time windows or failure to receive a dose. For example, if an individual received the first dose at 30 days after birth (Patient A in **Figure 1**), both clones would be censored at day 30 under the recommended and delayed schedules due to the early timing of the first dose. If an individual received the first dose at 42 days and the second dose at 70 days after birth (Patient B in **Figure 1**), its clone assigned to the recommended schedule would be followed until the end of the two-year follow-up period, whereas the clone assigned to the delayed schedule would be censored at day 42 because the first dose was administered outside the delayed timing window.

To account for informative censoring, inverse probability of censoring weights (IPCW) were estimated from a Cox proportional hazards (PH) model under both dosing schedules, adjusted for infant sex, birth weight z-score, birth height z-score, and the WAMI index for the outcome of being censored.^24,28,29^ Using the cloned and weighted dataset, we estimated the counterfactual cumulative risk of rotavirus infection and diarrhea under scenarios in which the target population followed either the recommended or delayed schedule in Brazil and Peru. We computed 95% confidence intervals (CIs) through a nonparametric bootstrap with 200 resamples.^30,31^ Differences between the recommended and delayed schedules were summarized using weighted cumulative risk ratios (RR) with the risk under the recommended schedule as a reference.

All analyses were conducted with R (R Center for Statistical Computing; Vienna, Austria) v4.2.1. Censoring weights were estimated using the ‘survival’ package.^32^ R scripts can be found at the following GitHub repository: https://github.com/KayokoShioda/DOSETTE_RotaV_MALED.

### Sensitivity analysis

Because the sample size in each country in the MAL-ED study was not large, we repeated the aforementioned clone-censor weight analysis with a combined study population of Peru and Brazil (n=346). We adjusted for country in IPCW calculation in the Cox PH model.

## Results

### Dosing schedules and rotavirus outcomes in Brazil and Peru

After excluding children with missing data, 192 children in Peru and 154 children in Brazil were included in the analysis. Of these, 190 (99%) children in Peru and 119 (77.4%) in Brazil received two doses of the rotavirus vaccine (**Table 1**). In Peru, 159 (82.8%) children received the vaccine according to the recommended schedule, while 28 (14.6%) received it according to the delayed schedule. Similarly, in Brazil, 66 (42.9%) children received the vaccine according to the recommended schedule, while 51 (33.1%) received it according to the delayed schedule.

**Table 1.** Characteristics of children included in the target trial emulation.

| Characteristics | Peru (N=192) | Brazil (N=154) |
| --- | --- | --- |
| Sex [n (%)] |  |  |
| Female | 87 (45.3%) | 71 (46.1%) |
| Male | 105 (54.7%) | 84 (54.6%) |
| Age at first dose (days) [mean (SD)] | 64.2 (8.3) | 70.8 (18.2) |
| Height at Birth z-score [mean (SD)] | -1.0 (1.0) | -0.8 (1.2) |
| Weight at Birth z-score [mean (SD)] | -0.6 (0.9) | -0.2 (1.1) |
| WAMI index [mean (SD)] | 0.6 (0.1) | 0.8 (0.08) |
| Number of rotavirus vaccine doses |  |  |
| Two [n (%)] | 190 (99.0%) | 119 (77.4%) |
| One [n (%)] | 1 (0.5%) | 26 (16.9%) |
| Zero [n (%)] | 1 (0.5%) | 9 (5.8%) |
| Timing of two-dose rotavirus vaccine [n (%)] |  |  |
| Recommended timing [n (%)] | 159 (82.8%) | 66 (42.9%) |
| Delayed timing [n (%)] | 28 (14.6%) | 51 (33.1%) |
| Rotavirus infection during two-year follow-up |  |  |
| At least one PCR-confirmed infection [n (%)] | 135 (70.3%) | 35 (22.7%) |
| At least one EIA-confirmed infection [n (%)] | 100 (52.1%) | 14 (9.1%) |
| At least one EIA-confirmed diarrhea [n (%)] | 65 (33.9%) | 4 (2.6%) |

Rotavirus incidence was higher in Peru than in Brazil. In Peru, 135 (70.3%) children experienced at least one PCR-confirmed rotavirus infection, 100 (52.1%) had EIA-confirmed infection, and 65 (33.9%) had EIA-confirmed rotavirus diarrhea; the corresponding proportions in Brazil were 35 (22.7%), 14 (9.1%), and 4 (2.6%), respectively (**Table 1**).

### Estimated cumulative risk of rotavirus outcomes under two different dosing schedules

We evaluated the rotavirus vaccine effectiveness against two outcomes: infection and diarrhea. The weighted cumulative risk of rotavirus infection among children under two years of age was similar under the recommended and delayed schedules in both countries. For PCR-confirmed rotavirus infection, the two-year cumulative risk was 0.70 (95% CI: 0.63-0.78) under the recommended schedule and 0.71 (0.52-0.94) under the delayed schedule in Peru (RR at two years: 1.0 [0.73-1.37]) (**Figure 2a and 2d**). The incidence of rotavirus infection was lower in Brazil, but the two-year cumulative risk remained similar between the recommended (0.19 [95% CI: 0.11-0.31]) and delayed schedules (0.16 [0.08-0.27]; RR at two years: 0.85 [0.31-1.66]) (**Figure 3a and 3d**). Similarly, the risk of EIA-confirmed rotavirus infection was similar between the recommended and delayed schedules both in Peru (RR at two years: 1.31 [95% CI: 0.87-2.04], **Figure 2b and 2e**) and in Brazil (1.00 [0.09-5.57], **Figure 3b and 3e**).

**Figure 2:**
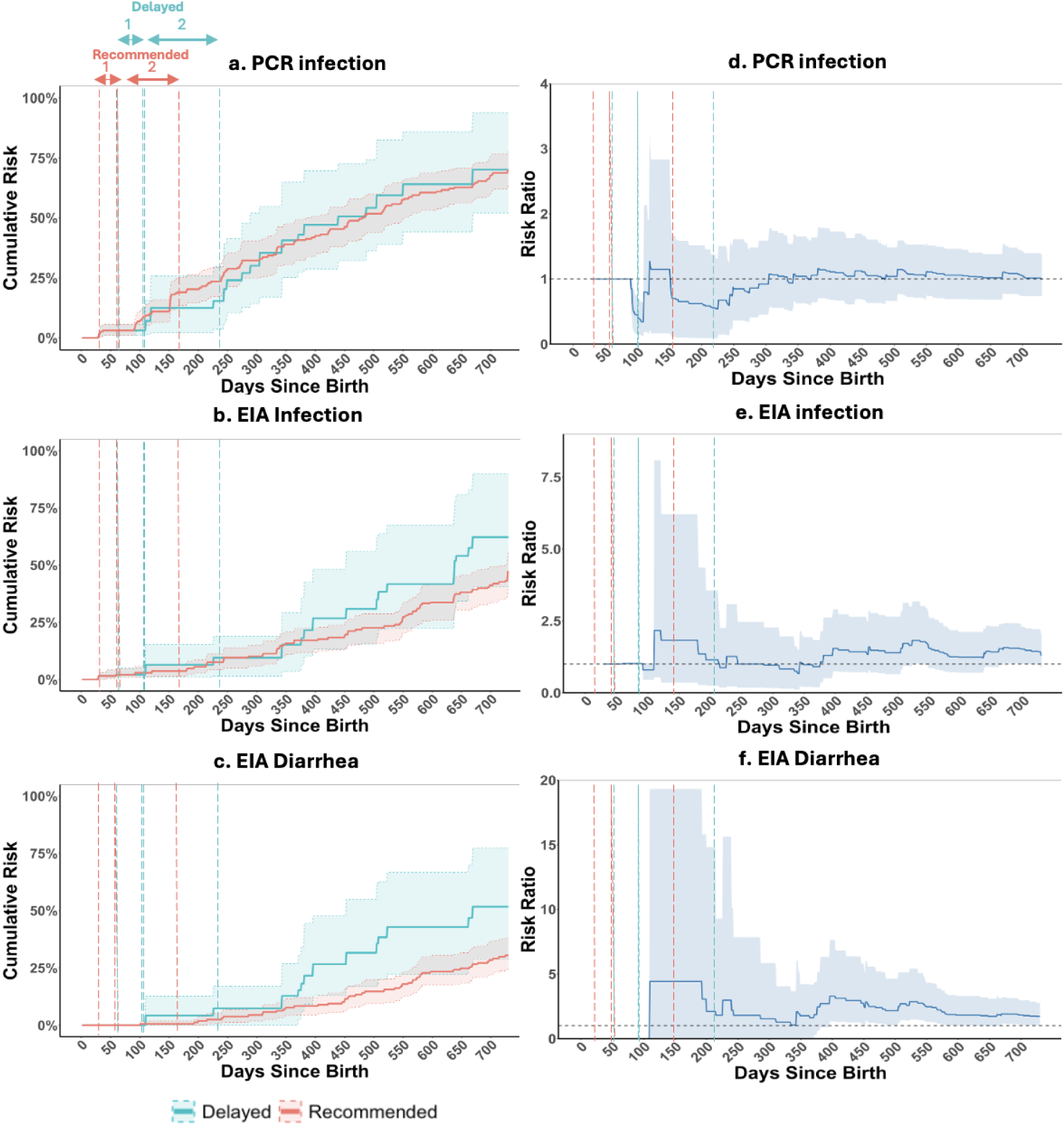
Estimated cumulative risks and risk ratios for rotavirus outcomes under recommended and delayed rotavirus vaccine dosing schedules in Peru. Panels on the left show the estimated cumulative risks of PCR-confirmed infection (panel a), EIA-confirmed infection (panel b), and EIA-confirmed rotavirus diarrhea (panel c) under the recommended (red) and delayed (green) dosing schedules during 730 days of follow-up in Peru. Panels on the right (panels d–f) show the corresponding risk ratios (the recommended schedule as a reference). Solid lines represent the point estimates, and shaded areas represent the 95% confidence intervals. Dashed vertical lines indicate the recommended (red) and delayed (green) time windows for the first and second vaccine doses.

**Figure 3:**
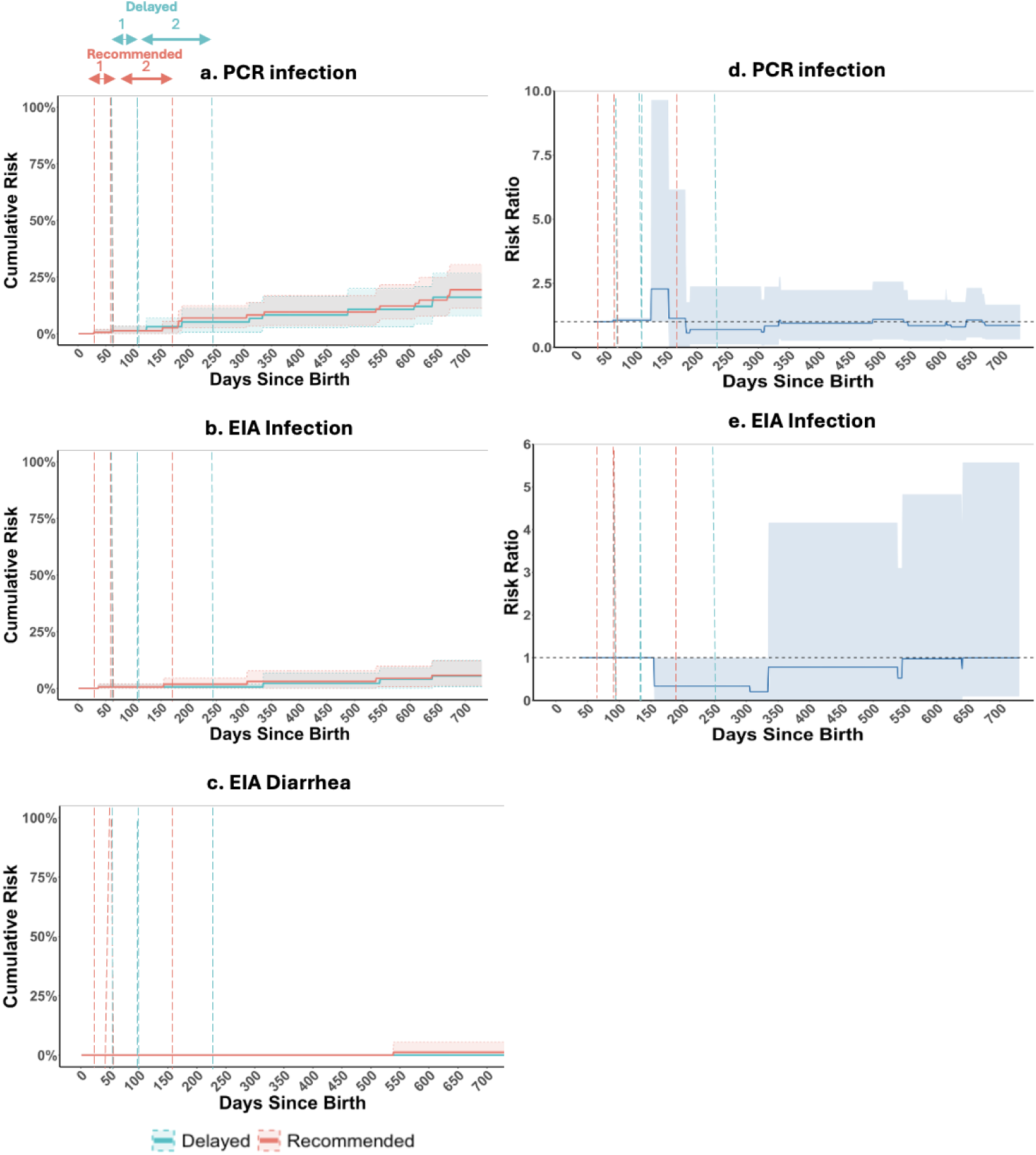
Estimated cumulative risks and risk ratios for rotavirus outcomes under recommended and delayed rotavirus vaccine dosing schedules in Brazil. Panels on the left show the estimated cumulative risks of PCR-confirmed infection (panel a), EIA-confirmed infection (panel b), and EIA-confirmed rotavirus diarrhea (panel c) under the recommended (red) and delayed (green) dosing schedules during 730 days of follow-up in Brazil. Panels on the right (panels d and e) show the corresponding risk ratios (the recommended schedule as a reference). Solid lines represent the point estimates, and shaded areas represent the 95% confidence intervals. Dashed vertical lines indicate the recommended (red) and delayed (green) time windows for the first and second vaccine doses. Risk ratios for EIA-confirmed rotavirus diarrhea are not shown because there were too few events to estimate stable risk ratios. Only four children experienced EIA-confirmed rotavirus diarrhea, resulting in estimated cumulative risks under the recommended schedule (the denominator of the risk ratio) that were zero or near zero during much of follow-up.

For EIA-confirmed rotavirus diarrhea, the delayed schedule yielded a higher cumulative risk compared to the recommended schedule in Peru, especially in the second year of life (RR at two years: 1.73 [1.02-2.77]) (**Figure 2c and 2e**). No difference was observed in Brazil, where only four children (2.6%) experienced rotavirus diarrhea (**Figure 3c**).

### Sensitivity Analysis

The trend of the estimated cumulative risk of various rotavirus outcomes remained similar between the recommended and delayed schedules after combining the data from Peru and Brazil (**Supplementary Figure 1**). The risk of rotavirus infections confirmed by PCR or EIA was similar between the two schedules (RR for PCR-confirmed infection at two years: 1.03 [95% CI: 0.74-1.49]; RR for EIA-confirmed infection at two years: 1.23 [0.79-1.91]), while the risk of EIA-confirmed rotavirus diarrhea was higher under the delayed schedule (RR at two years: 1.64 [0.92-2.61]).

## Discussion

This target trial emulation study compared the cumulative risk of rotavirus infection and diarrhea under two dosing schedules of the 2-dose rotavirus vaccine RV1 (recommended and delayed) among children under two years of age in Peru and Brazil. By estimating the risk of various rotavirus outcomes from birth through the two-year follow-up period, we assessed how delaying both doses of the RV1 vaccine may affect the real-world effectiveness in a high-transmission setting (Peru) and a low-transmission setting (Brazil). We found that the delayed schedule was associated with a higher cumulative risk of EIA-confirmed rotavirus diarrhea in Peru, while no difference was observed in Brazil due to the small number of children with rotavirus diarrhea. In contrast, the cumulative risk of rotavirus infection was similar under both schedules in both countries. This finding is consistent with the primary role of rotavirus vaccines in preventing diarrheal disease rather than infection.^33^ Although trials^19,34^ and immunogenicity studies^35,36^ have tested delayed rotavirus vaccine doses, to our knowledge, this is the first study to compare different timings of rotavirus vaccine doses against rotavirus infection and diarrhea in real-world settings.

The timing of rotavirus vaccination has been an area of ongoing investigation, particularly in LMICs where vaccine effectiveness is often lower than in high-income settings.^3–7^ There has been a hypothesis that delayed dosing schedules may improve immunogenicity and protection by administering doses at older ages when maternal antibody interference may be reduced and the infant’s immune system is more mature.^10,34^ One immunogenicity study among infants in Pakistan found that the antibody response after delayed administration of the 2-dose rotavirus vaccine at 10 and 14 weeks did not differ from that of the 6- and 10-week schedule.^36^ The other immunogenicity study among infants in rural Ghana observed an increase in the frequency of seroconversion after a delayed schedule at 10 and 14 weeks of age, compared to the 6- and 10-week schedule, though a difference was not significant.^35^ One RCT in Bangladesh suggested the delayed schedule at 10 and 17 weeks may improve immune protection against severe rotavirus infection, while this trial did not directly compare the delayed schedule to the recommended schedule.^19^ Our study adds another finding, suggesting that delaying vaccination does not change the risk of infection (including both symptomatic and asymptomatic ones) in the first two years of life. In contrast, for symptomatic infection, the risk was higher under the delayed schedule in one of the two countries (Peru) in our analysis. This risk remained similar between schedules in Brazil, which may be due to the small number of children that experienced the outcome. Future studies with larger sample sizes may be beneficial to evaluate the risk of symptomatic infection under more granular dose-timing schedules.

Target trial emulation has increasingly been applied to vaccine evaluation, particularly following the COVID-19 pandemic.^37^ A scoping review showed a growing number of studies using this approach to evaluate vaccine effectiveness, including the number and timing of doses, across vaccines such as COVID-19, rotavirus, mpox, and influenza.^37^ While most applications have been done in high-income settings^37^, this study contributes to the literature by applying target trial emulation to vaccine evaluation in non-high-income settings (Brazil and Peru).

It is important to note that this study focused on comparing the direct effect of the two dosing schedules. Many vaccines also have population-level effects by reducing transmission, thereby lowering the risk of infection among both vaccinated and unvaccinated individuals. These indirect effects and changes in transmission dynamics were not part of the evaluation in this study. Evaluating such effects would require data from multiple populations with varying levels of vaccine coverage. Extending target trial emulation frameworks to incorporate these population-level dynamics is an important direction for future methodological research.

This study has several limitations. First, our samples may not be representative of the general pediatric populations of Peru and Brazil. As MAL-ED participants were regularly followed by study staff, they may have been more adherent to vaccine schedules than the broader population. Second, the sample sizes in Peru and Brazil were relatively small. There were 159 children in Peru who followed the recommended dosing schedule, and 28 who followed the delayed dosing schedule. In Brazil, 66 followed the recommended protocol while 51 followed the delayed protocol. These small subgroups limit the statistical power to detect small or moderate differences in the cumulative infection risk between the two protocols, especially when outcome incidence is low.

In conclusion, using observational data from a multi-country birth cohort, we emulated a target trial to compare the direct effects of recommended and delayed timing of a 2-dose rotavirus vaccine. In both a high-incidence setting (Peru) and a low-incidence setting (Brazil), the two schedules yielded similar risks of infection. Our analysis suggested that the delayed schedule may increase the risk of diarrhea, but further analysis would be necessary to understand if this increase is significant and generalizable.

## Data Availability

All data used in the present study are available upon reasonable request to the original data owners

## Acknowledgement

This study was made possible by cooperative agreement CDC-RFA-FT-23-0069 from the CDC’s Center for Forecasting and Outbreak Analytics. The funders of the study had no role in study design, data collection, data analysis, data interpretation, or writing of the report. Its contents are solely the responsibility of the authors and do not necessarily represent the official views of the funders. K.S. and S.G. were also supported by the Center for Health Data Science at Boston University.

## Supplementary Methods

### Data from the MAL-ED Study

In Peru, the vaccination dataset included 261 participants, and the infection dataset included 194 children, of which 192 of those had both vaccination and infection data (69 had no infection data, and two had no vaccination data). Thus, there were a total of 192 participants from Peru.

In Brazil, the vaccination dataset included 186 children, and the infection dataset included 165 children, of which 155 of those participants had both vaccination and infection data (31 had no infection data, and 10 had no vaccination data). One participant in Brazil had a data entry error showing that they received dose 1 of RV1 on their day of birth so they were excluded from the analysis. The total study population in Brazil was 154 participants.

## Supplementary Tables

**Supplementary Table 1:**
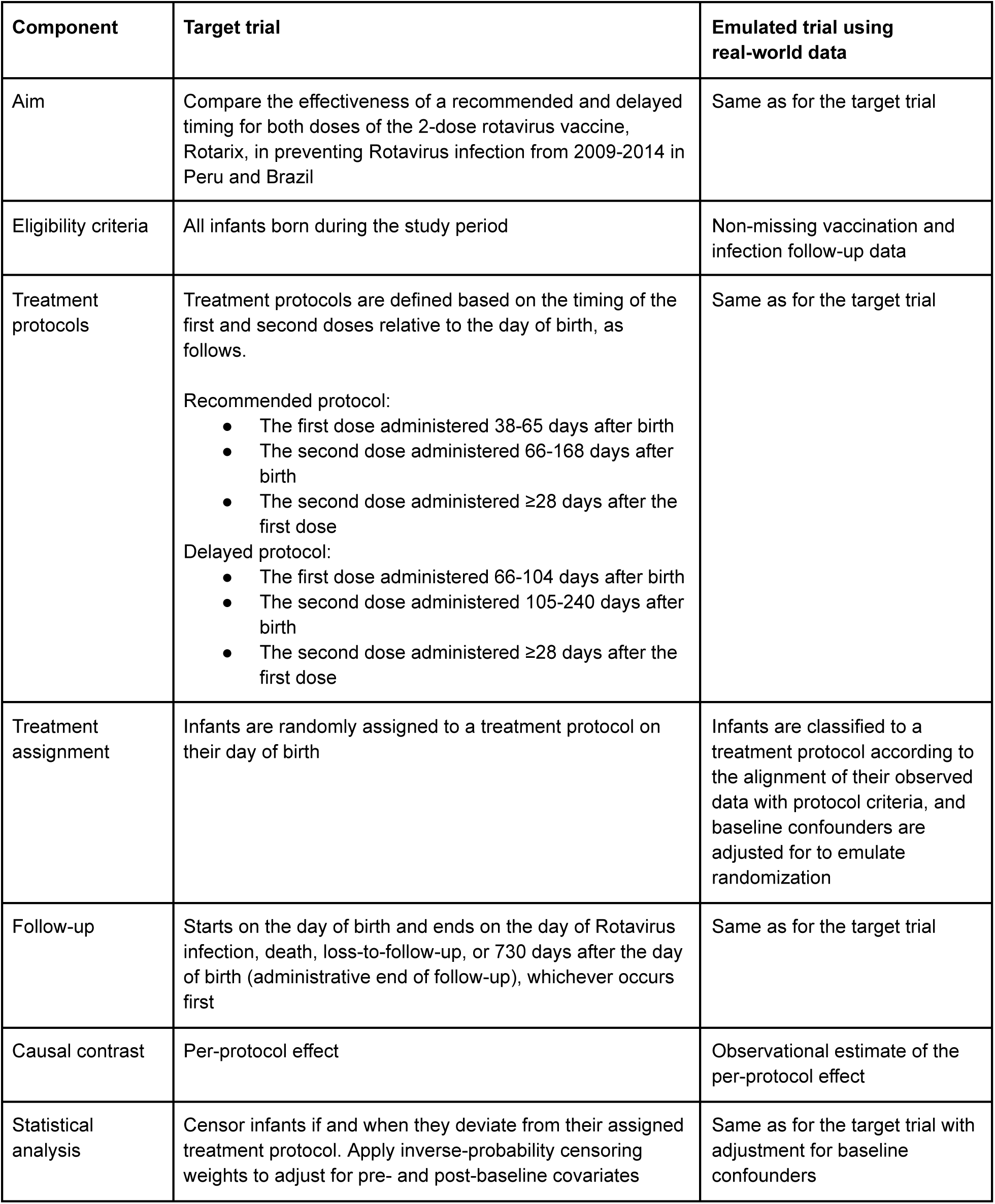
Specification of target trial emulation from hypothetical target trial.

| Component | Target trial | Emulated trial using real-world data |
| --- | --- | --- |
| Aim | Compare the effectiveness of a recommended and delayed timing for both doses of the 2-dose rotavirus vaccine, Rotarix, in preventing Rotavirus infection from 2009-2014 in Peru and Brazil | Same as for the target trial |
| Eligibility criteria | All infants born during the study period | Non-missing vaccination and infection follow-up data |
| Treatment protocols | Treatment protocols are defined based on the timing of the first and second doses relative to the day of birth, as follows.<br>Recommended protocol:<br>• The first dose administered 38-65 days after birth<br>• The second dose administered 66-168 days after birth<br>• The second dose administered $\geq 28$ days after the first dose<br>Delayed protocol:<br>• The first dose administered 66-104 days after birth<br>• The second dose administered 105-240 days after birth<br>• The second dose administered $\geq 28$ days after the first dose | Same as for the target trial |
| Treatment assignment | Infants are randomly assigned to a treatment protocol on their day of birth | Infants are classified to a treatment protocol according to the alignment of their observed data with protocol criteria, and baseline confounders are adjusted for to emulate randomization |
| Follow-up | Starts on the day of birth and ends on the day of Rotavirus infection, death, loss-to-follow-up, or 730 days after the day of birth (administrative end of follow-up), whichever occurs first | Same as for the target trial |
| Causal contrast | Per-protocol effect | Observational estimate of the per-protocol effect |
| Statistical analysis | Censor infants if and when they deviate from their assigned treatment protocol. Apply inverse-probability censoring weights to adjust for pre- and post-baseline covariates | Same as for the target trial with adjustment for baseline confounders |

**Supplementary Table 2:**
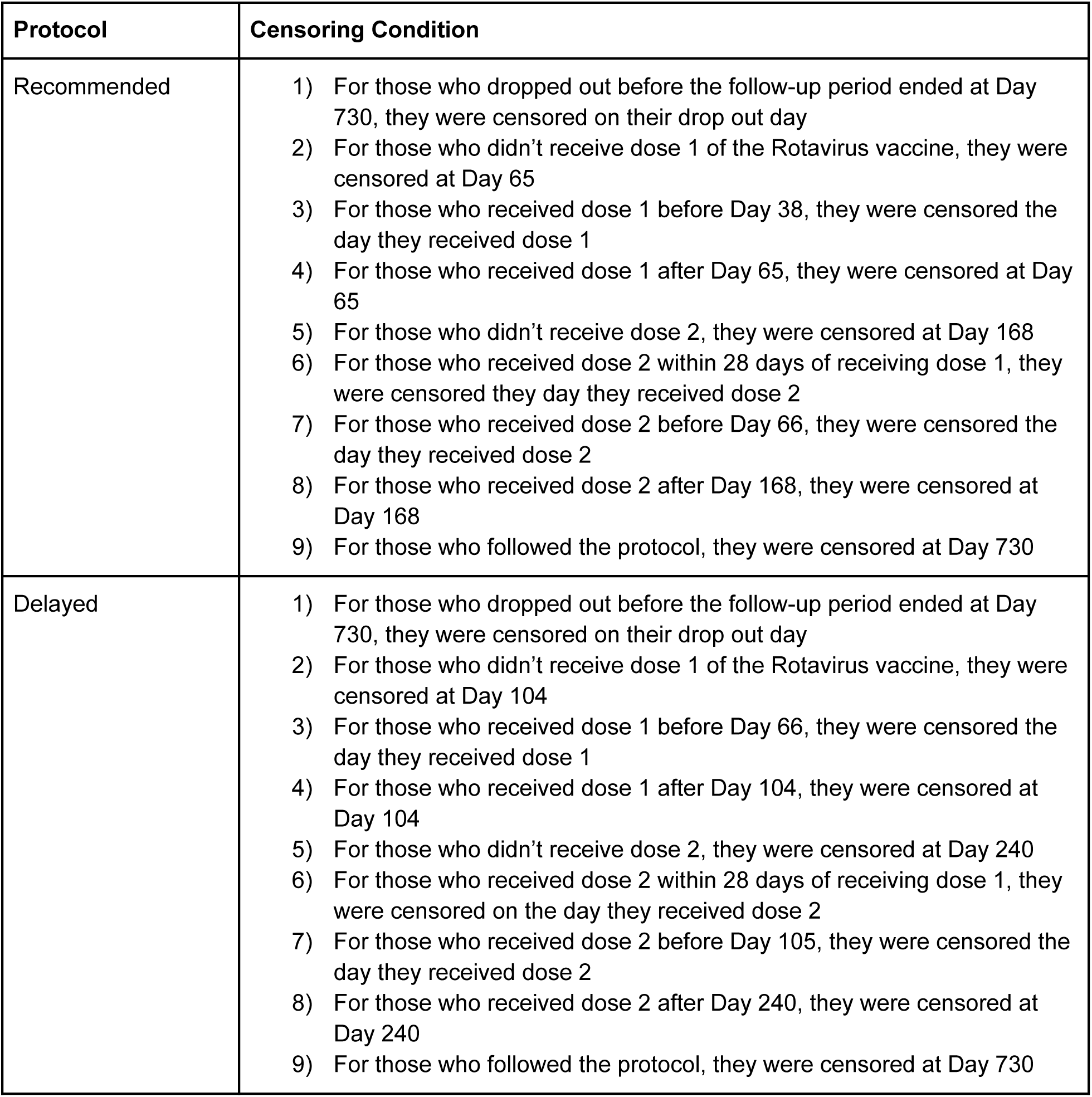
Conditions for censoring corresponding to the Rotavirus vaccine protocols.

## Supplementary Figures

**Supplementary Figure 1:**
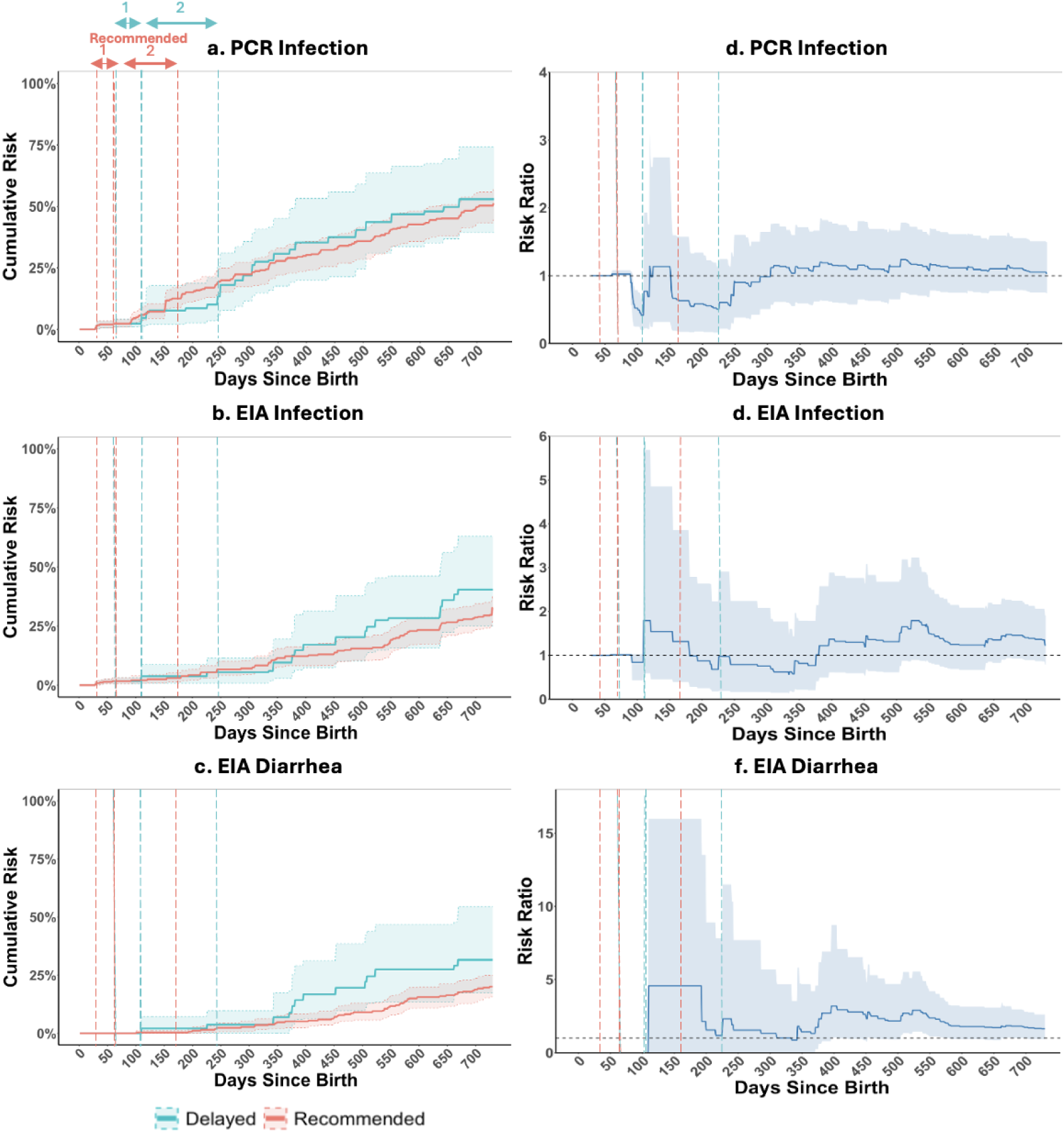
Estimated cumulative risks and risk ratios for rotavirus outcomes under recommended and delayed rotavirus vaccine dosing schedules in Peru and Brazil. Panels on the left show the estimated cumulative risks of PCR-confirmed infection (panel a), EIA-confirmed infection (panel b), and EIA-confirmed rotavirus diarrhea (panel c) under the recommended (red) and delayed (green) dosing schedules during 730 days of follow-up in Peru. Panels on the right (panels d–f) show the corresponding risk ratios (the recommended schedule as a reference). Solid lines represent the point estimates, and shaded areas represent the 95% confidence intervals. Dashed vertical lines indicate the recommended and delayed time windows for the first and second vaccine doses.

## References

1. International Vaccine Access Center | Johns Hopkins Bloomberg School of Public Health [Internet]. [cited 2026 Apr 19]. Available from: https://publichealth.jhu.edu/ivac

2. Clark A, Mahmud S, Debellut F, Pecenka C, Jit M, Perin J, et al. Estimating the global impact of rotavirus vaccines on child mortality. Int J Infect Dis. 2023 Dec;137:90–7. doi:10.1016/j.ijid.2023.10.005 PubMed PMID: 37863311; PubMed Central PMCID: PMC10689250.

3. Bhandari N, Rongsen-Chandola T, Bavdekar A, John J, Antony K, Taneja S, et al. Efficacy of a monovalent human-bovine (116E) rotavirus vaccine in Indian infants: a randomised, double-blind, placebo-controlled trial. Lancet. 2014 Jun 21;383(9935):2136–43. doi:10.1016/S0140-6736(13)62630-6 PubMed PMID: 24629994; PubMed Central PMCID: PMC4532697.

4. Cunliffe NA, Witte D, Ngwira BM, Todd S, Bostock NJ, Turner AM, et al. Efficacy of human rotavirus vaccine against severe gastroenteritis in Malawian children in the first two years of life: a randomized, double-blind, placebo controlled trial. Vaccine. 2012 Apr 27;30 Suppl 1(0 1):A36–43. doi:10.1016/j.vaccine.2011.09.120 PubMed PMID: 22520135; PubMed Central PMCID: PMC3982044.

5. Clark A, van Zandvoort K, Flasche S, Sanderson C, Bines J, Tate J, et al. Efficacy of live oral rotavirus vaccines by duration of follow-up: a meta-regression of randomised controlled trials. Lancet Infect Dis. 2019 Jul;19(7):717–27. doi:10.1016/S1473-3099(19)30126-4 PubMed PMID: 31178289; PubMed Central PMCID: PMC6595176.

6. Parker EP, Ramani S, Lopman BA, Church JA, Iturriza-Gómara M, Prendergast AJ, et al. Causes of impaired oral vaccine efficacy in developing countries. Future Microbiol. 2018 Jan;13(1):97–118. doi:10.2217/fmb-2017-0128 PubMed PMID: 29218997; PubMed Central PMCID: PMC7026772.

7. Ramakrishnan G, Ma JZ, Haque R, Petri WA. Rotavirus vaccine protection in low-income and middle-income countries. The Lancet Infectious Diseases. 2019 Jul 1;19(7):673–4. doi:10.1016/S1473-3099(19)30263-4 PubMed PMID: 31178290.

8. Vesikari T, Matson DO, Dennehy P, Van Damme P, Santosham M, Rodriguez Z, et al. Safety and efficacy of a pentavalent human-bovine (WC3) reassortant rotavirus vaccine. N Engl J Med. 2006 Jan 5;354(1):23–33. doi:10.1056/NEJMoa052664 PubMed PMID: 16394299.

9. Bhandari N, Rongsen-Chandola T, Bavdekar A, John J, Antony K, Taneja S, et al. Efficacy of a monovalent human-bovine (116E) rotavirus vaccine in Indian children in the second year of life. Vaccine. 2014 Aug 11;Rotavirus in India: An update on epidemiology and vaccines 32:A110–6. doi:10.1016/j.vaccine.2014.04.079

10. Lopman BA, Pitzer VE, Sarkar R, Gladstone B, Patel M, Glasser J, et al. Understanding Reduced Rotavirus Vaccine Efficacy in Low Socio-Economic Settings. PLOS ONE. 2012 Aug 6;7(8):e41720. doi:10.1371/journal.pone.0041720

11. Diarrhoeal disease [Internet]. [cited 2026 Jun 16]. Available from: https://www.who.int/news-room/fact-sheets/detail/diarrhoeal-disease

12. Black RE, Perin J, Yeung D, Rajeev T, Miller J, Elwood SE, et al. Estimated global and regional causes of deaths from diarrhoea in children younger than 5 years during 2000-21: a systematic review and Bayesian multinomial analysis. Lancet Glob Health. 2024 Jun;12(6):e919–28. doi:10.1016/S2214-109X(24)00078-0 PubMed PMID: 38648812; PubMed Central PMCID: PMC11099298.

13. El Adam S, Zou M, Kim S, Henry B, Krajden M, Skowronski DM. SARS-CoV-2 mRNA Vaccine Effectiveness in Health Care Workers by Dosing Interval and Time Since Vaccination: Test-Negative Design, British Columbia, Canada. Open Forum Infect Dis. 2022 May;9(5):ofac178. doi:10.1093/ofid/ofac178 PubMed PMID: 35531384; PubMed Central PMCID: PMC9047244.

14. Le Vu S, Bertrand M, Semenzato L, Jabagi MJ, Botton J, Drouin J, et al. Influence of mRNA Covid-19 vaccine dosing interval on the risk of myocarditis. Nat Commun. 2024 Sep 5;15(1):7745. doi:10.1038/s41467-024-52038-6

15. Buchan SA, Seo CY, Johnson C, Alley S, Kwong JC, Nasreen S, et al. Epidemiology of Myocarditis and Pericarditis Following mRNA Vaccination by Vaccine Product, Schedule, and Interdose Interval Among Adolescents and Adults in Ontario, Canada. JAMA Netw Open. 2022 Jun 1;5(6):e2218505. doi:10.1001/jamanetworkopen.2022.18505 PubMed PMID: 35749115; PubMed Central PMCID: PMC9233237.

16. Shioda K, Breskin A, Harati P, Chamberlain AT, Komura T, Lopman BA, et al. Comparative effectiveness of alternative intervals between first and second doses of the mRNA COVID-19 vaccines. Nat Commun. 2024 Feb 9;15(1):1214. doi:10.1038/s41467-024-45334-8

17. Rodrigues CMC, Plotkin SA. The influence of interval between doses on response to vaccines. Vaccine. 2021 Dec 3;39(49):7123–7. doi:10.1016/j.vaccine.2021.10.050 PubMed PMID: 34774357; PubMed Central PMCID: PMC8580840.

18. Payne RP, Longet S, Austin JA, Skelly DT, Dejnirattisai W, Adele S, et al. Immunogenicity of standard and extended dosing intervals of BNT162b2 mRNA vaccine. Cell. 2021 Nov 11;184(23):5699–5714.e11. doi:10.1016/j.cell.2021.10.011 PubMed PMID: 34735795; PubMed Central PMCID: PMC8519781.

19. Colgate ER, Haque R, Dickson DM, Carmolli MP, Mychaleckyj JC, Nayak U, et al. Delayed Dosing of Oral Rotavirus Vaccine Demonstrates Decreased Risk of Rotavirus Gastroenteritis Associated With Serum Zinc: A Randomized Controlled Trial. Clin Infect Dis. 2016 Sep 1;63(5):634–41. doi:10.1093/cid/ciw346 PubMed PMID: 27217217.

20. Skowronski DM, Febriani Y, Ouakki M, Setayeshgar S, El Adam S, Zou M, et al. Two-Dose Severe Acute Respiratory Syndrome Coronavirus 2 Vaccine Effectiveness With Mixed Schedules and Extended Dosing Intervals: Test-Negative Design Studies From British Columbia and Quebec, Canada. Clin Infect Dis. 2022 Dec 1;75(11):1980–92. doi:10.1093/cid/ciac290

21. Martinez DR, Ooi EE. A potential silver lining of delaying the second dose. Nat Immunol. 2022 Mar;23(3):349–51. doi:10.1038/s41590-022-01143-z

22. Immunization Data [Internet]. [cited 2026 Apr 19]. WHO Immunization Data portal - Detail Page. Available from: https://immunizationdata.who.int/global/wiise-detail-page

23. Zaman K, Dang DA, Victor JC, Shin S, Yunus M, Dallas MJ, et al. Efficacy of pentavalent rotavirus vaccine against severe rotavirus gastroenteritis in infants in developing countries in Asia: a randomised, double-blind, placebo-controlled trial. Lancet. 2010 Aug 21;376(9741):615–23. doi:10.1016/S0140-6736(10)60755-6 PubMed PMID: 20692031.

24. Hernán MA, Robins JM. Using Big Data to Emulate a Target Trial When a Randomized Trial Is Not Available. Am J Epidemiol. 2016 Apr 15;183(8):758–64. doi:10.1093/aje/kwv254 PubMed PMID: 26994063; PubMed Central PMCID: PMC4832051.

25. Etiology, Risk Factors and Interactions of Enteric Infections and Malnutrition and the Consequences for Child Health and Development (MAL-ED). FNIH [Internet]. [cited 2026 Apr 19]. Available from: https://fnih.org/our-programs/etiology-risk-factors-and-interactions-of-enteric-infections-and-m alnutrition-and-the-consequences-for-child-health-and-development-mal-ed/

26. Seheri LM, Page NA, Mawela MPB, Mphahlele MJ, Steele AD. Rotavirus vaccination within the South African Expanded Programme on Immunisation. Vaccine. 2012 Sep 7;Introducing New Vaccines into the South African National Immunisation Programme - a Case Study 30:C14–20. doi:10.1016/j.vaccine.2012.04.018

27. Richard SA, McCormick BJ, Murray-Kolb LE, Lee GO, Seidman JC, Mahfuz M, et al. Enteric dysfunction and other factors associated with attained size at 5 years: MAL-ED birth cohort study findings. The American Journal of Clinical Nutrition. 2019 Jul 1;110(1):131–8. doi:10.1093/ajcn/nqz004

28. Maringe C, Benitez Majano S, Exarchakou A, Smith M, Rachet B, Belot A, et al. Reflection on modern methods: trial emulation in the presence of immortal-time bias. Assessing the benefit of major surgery for elderly lung cancer patients using observational data. Int J Epidemiol. 2020 Oct 1;49(5):1719–29. doi:10.1093/ije/dyaa057 PubMed PMID: 32386426; PubMed Central PMCID: PMC7823243.

29. Hernán MA, Sauer BC, Hernández-Díaz S, Platt R, Shrier I. Specifying a target trial prevents immortal time bias and other self-inflicted injuries in observational analyses. J Clin Epidemiol. 2016 Nov;79:70–5. doi:10.1016/j.jclinepi.2016.04.014 PubMed PMID: 27237061; PubMed Central PMCID: PMC5124536.

30. Efron B, Tibshirani RJ. An Introduction to the Bootstrap. New York: Chapman and Hall/CRC; 1994. 456 p. doi:10.1201/9780429246593

31. Sherman M, Cessie SL. A comparison between bootstrap methods and generalized estimating equations for correlated outcomes in generalized linear models. Communications in Statistics - Simulation and Computation. 1997 Jan;26(3):901–25. doi:10.1080/03610919708813417

32. Therneau TM. survival: Survival Analysis [Internet]. 2001 [cited 2026 Apr 19]. p. 3.8–6. Available from: https://CRAN.R-project.org/package=survivaldoi:10.32614/CRAN.package.survival

33. Cates JE, Tate JE, Parashar U. Rotavirus vaccines: progress and new developments. Expert Opinion on Biological Therapy. 2022 Mar 4;22(3):423–32. doi:10.1080/14712598.2021.1977279 PubMed PMID: 34482790.

34. Madhi SA, Cunliffe NA, Steele D, Witte D, Kirsten M, Louw C, et al. Effect of human rotavirus vaccine on severe diarrhea in African infants. N Engl J Med. 2010 Jan 28;362(4):289–98. doi:10.1056/NEJMoa0904797 PubMed PMID: 20107214.

35. Armah G, Lewis KDC, Cortese MM, Parashar UD, Ansah A, Gazley L, et al. A Randomized, Controlled Trial of the Impact of Alternative Dosing Schedules on the Immune Response to Human Rotavirus Vaccine in Rural Ghanaian Infants. J Infect Dis. 2016 Jun 1;213(11):1678–85. doi:10.1093/infdis/jiw023 PubMed PMID: 26823335; PubMed Central PMCID: PMC4857471.

36. Ali SA, Kazi AM, Cortese MM, Fleming JA, Parashar UD, Jiang B, et al. Impact of different dosing schedules on the immunogenicity of the human rotavirus vaccine in infants in Pakistan: a randomized trial. J Infect Dis. 2014 Dec 1;210(11):1772–9. doi:10.1093/infdis/jiu335 PubMed PMID: 24939906.

37. Komura T, Watanabe M, Shioda K. Exploring the application of target trial emulation in vaccine evaluation: scoping review. Am J Epidemiol. 2025 Oct 7;194(10):3028–40. doi:10.1093/aje/kwaf053 PubMed PMID: 40069950.

